# Association between overcrowded households, multigenerational households, and COVID-19: a cohort study

**DOI:** 10.1101/2021.06.14.21258904

**Authors:** Arnab K. Ghosh, Sara Venkatraman, Orysya Soroka, Evgeniya Reshetnyak, Mangala Rajan, Anjile An, John K. Chae, Christopher Gonzalez, Jonathan Prince, Charles DiMaggio, Said Ibrahim, Monika M. Safford, Nathaniel Hupert

## Abstract

**Introduction:** The role of overcrowded and multigenerational households as a risk factor for COVID-19 remains unmeasured. The objective of this study is to examine and quantify the association between overcrowded and multigenerational households, and COVID-19 in New York City (NYC).

**Methods:** We conducted a Bayesian ecological time series analysis at the ZIP Code Tabulation Area (ZCTA) level in NYC to assess whether ZCTAs with higher proportions of overcrowded (defined as proportion of estimated number of housing units with more than one occupant per room) and multigenerational households (defined as the estimated percentage of residences occupied by a grandparent and a grandchild less than 18 years of age) were independently associated with higher suspected COVID-19 case rates (from NYC Department of Health Syndromic Surveillance data for March 1 to 30, 2020). Our main measure was adjusted incidence rate ratio (IRR) of suspected COVID-19 cases per 10,000 population. Our final model controlled for ZCTA-level sociodemographic factors (median income, poverty status, White race, essential workers), prevalence of clinical conditions related to COVID-19 severity (obesity, hypertension, coronary heart disease, diabetes, asthma, smoking status, and chronic obstructive pulmonary disease), and spatial clustering.

**Results:** 39,923 suspected COVID-19 cases presented to emergency departments across 173 ZCTAs in NYC. Adjusted COVID-19 case rates increased by 67% (IRR 1.67, 95% CI = 1.12, 2.52) in ZCTAs in quartile four (versus one) for percent overcrowdedness and increased by 77% (IRR 1.77, 95% CI = 1.11, 2.79) in quartile four (versus one) for percent living in multigenerational housing. Interaction between both exposures was not significant (β_interaction_ = 0.99, 95% CI: 0.99-1.00).

**Conclusions:** Over-crowdedness and multigenerational housing are independent risk factors for suspected COVID-19. In the early phase of surge in COVID cases, social distancing measures that increase house-bound populations may inadvertently but temporarily increase SARS-CoV-2 transmission risk and COVID-19 disease in these populations.

## Introduction

The COVID-19 pandemic has exposed striking health-related disparities in minority populations in the United States. Studies have highlighted disparities in COVID-19 testing, morbidity, and mortality between non-Hispanic Whites, non-Hispanic Blacks, and Hispanic Americans in both health care^1,2^ and community settings.^3-6^ Existing well-described disparities in health status, access to healthcare, and other social determinants may account for these differences. Furthermore, a growing body of evidence suggests that socioeconomic factors (such as “essential worker” status) may play a major role in risk of acquiring SARS-CoV-2 infection and differential morbidity and mortality from COVID-19.^1,3,4^ To date, however, the role of overcrowding and household composition on transmission of SARS-CoV2 and diagnosis of COVID-19 remains poorly understood.

The evolving science informing the transmission dynamics of SARS-CoV2 suggests that both proximity to infected patients^7,8^ and concentration of inoculum^9^ may play an important role in both acquisition of infection and subsequent severity of illness.^10,11^ In particular, individuals residing in overcrowded and multigenerational households may be at increased risk of developing more severe forms of COVID-19,^12^ since these settings reduce personal space and increase the risk of multiple exposures to a high inoculum of SARS-CoV-2 infection.^11,13^

In this study, we investigated whether ZIP code tabulation areas (ZCTA) with a higher percentage of overcrowded or multigenerational households represented independent risk factors for severe COVID-19 after accounting for other area-level socioeconomic, clinical, and spatiotemporal factors. We defined severe COVID-19 as patients presenting to the emergency department with suspected COVID-19-like symptoms.

## Methods

### Study Setting

We conducted a retrospective ZCTA-level time series analysis of daily suspected emergency department-presenting COVID-19 cases per 10,000 total population in each ZCTA in NYC in March 2020. Beginning in late February 2020, NYC became the global epicenter of the COVID-19 pandemic, with a first wave that crested in early April 2020. We chose to limit our analysis to March 2020 because we hypothesized that the role of multigenerational households and household overcrowdedness would be greatest at the start of the first wave of the COVID-19 pandemic in NYC both before and after the imposition of social distancing measures.

### Variables and Data Sources

The dependent variable was the daily count by ZCTA of suspected COVID-19 cases presenting to one of the 53 hospital emergency departments in NYC. These data were obtained from the NYC Department of Health and Mental Hygiene (NYC DOHMH) Syndromic Surveillance system, which contains suspected COVID-19 cases for 173 ZCTAs. Suspected COVID-19 cases were defined as cases of pneumonia and influenza-like illness that appeared after the 2019-20 influenza season had ended (based on laboratory analysis, in February 2020).^14^ This study used surveillance-based suspected cases rather than test-positive COVID-19 cases for two reasons: first, there was potentially vast undertesting for COVID-19 early in the pandemic in New York City, and second, daily test-positive COVID-19 case counts by residential ZCTA were not available. All data were obtained up to March 30, 2020.

The two exposure variables included jointly in the model were: 1) percentage of overcrowded housing by ZCTA, defined as the estimated number of housing units with more than one occupant per room, divided by the number of occupied housing units; and 2) percentage of multigenerational housing by ZCTA, defined as the estimated percentage of residences occupied by a grandparent and a grandchild less than 18 years of age. Both were obtained from the American Community Survey 5-year 2018 estimates. Both exposures were segmented into quartiles, with the first quartile (representing the least overcrowded or lowest proportion of multigenerational households) as reference.

Several socioeconomic factors have been reported to be associated with a greater risk of COVID-19 infection and severity. These include factors related to lower socioeconomic status such as lower median income,^1^ minority status,^2^ lack of health insurance,^5^ and an individual’s role as an essential worker.^15^ To account for these associations in an ecological analysis, we controlled for ZCTA-level estimates of total population, percentage of patients living below the federal poverty line (FPL), median income, percentage of White residents, and percentage of essential workers by ZCTA. The percentage of essential workers by ZCTA was calculated by replicating a method employed by the NYC Office of the Comptroller.^15^ The proportion of essential workers in each ZCTA was identified from service-oriented non-public roles using Census Industrial Classification Codes in the following categories: 1) public transit workers; 2) grocery, convenience and drug store workers; 3) trucking, warehouse and postal service workers; 4) healthcare workers; 5) childcare, homeless, food and family service workers; and 6) building cleaning service workers.

The model also controlled for clinical factors identified by the U.S. Centers for Disease Control and Prevention (CDC) that increase the risk for developing more severe COVID-19 illness.^16^ ZCTA-level data on COVID-19 disease clinical risk factors were obtained and derived from the CDC 500 Cities dataset 2016-2017. These included the percentage of obese adults (defined as body mass index [BMI] ≥ 30 kg/m^2^), percentage of adults who smoke, and percentage of adults with coronary heart disease, hypertension, diabetes, asthma, or chronic obstructive pulmonary disease (COPD).

In order to undertake descriptive and inferential spatial analyses, spatial shapefiles of NYC’s ZCTAs were downloaded and derived from the New York City Department of City Planning.^4^

### Statistical Analysis

Visual depictions of the suspected severe COVID-19 cases by ZCTA, percent of overcrowded households, and percent of multigenerational households were created using QGIS software. We used Moran’s I^17^ to formally assess spatial clustering of suspected severe COVID-19 cases, for which the null hypothesis is that suspected COVID-19 cases were randomly dispersed across the city.

To evaluate the average effect of household overcrowdedness and multigenerational households on suspected COVID-19 cases within each ZCTA, we first used a generalized linear model specifying a Poisson distribution for the dependent variable (using quasi-likelihood estimation to account for overdispersion), and robust standard errors. The model included daily time fixed effects, and suspected case counts by ZCTA were interpolated to account for the end-of-week variation in cases. All models used the population of individuals residing in each ZCTA as the population offset, with results interpreted as incidence rate ratios (IRR). To assess the robustness of the percent overcrowdedness and percent multigenerational households within each ZCTA as independent risk factors, we added the aforementioned control variables, which reflected reported ZCTA-level socioeconomic associations with COVID-19 exposure and clinical risk of COVID-19 severity, to a baseline model that consisted of an intercept term, the two exposure variables, and time in days. Variance inflation factors (VIF) were used to assess for multi-collinearity, and covariates with VIF > 10 were dropped sequentially. The variables dropped from the final, adjusted analysis were prevalence of chronic obstructive pulmonary disease, diabetes, and hypertension at the ZCTA-level.

Upon finding that there was significant spatial autocorrelation in our dependent variable (Moran’s I: 0.456, p = 0.001), we then fit a Bayesian version of our quasi-Poisson model using the integrated nested Laplace approximation (INLA) method, a computationally efficient way of fitting models to data exhibiting spatial or temporal structure. This method, described by Blangiardo et al.,^18^ models the variation in the dependent variable using a spatially-structured random effects term that accounts for local geographic influences indicated by the Moran’s I. This random effects term follows a conditional autoregressive model, meaning that the random variation in a ZCTA’s suspected COVID-19 case counts is modeled as the mean of the random effect terms for the adjacent ZCTAs. Temporal structure is accounted for in the model by both fixed and random effects. The temporal random effect is modeled by a random walk, which assumes that a ZCTA’s suspected case count at time *t* in days is equal to the case count at time *t*-1 plus some amount of random noise that is normally distributed with mean 0.

The final, fully adjusted model used the INLA method and treated the number of suspected COVID-19 cases in each ZCTA as a Poisson-distributed variable. The model consisted of a population offset; the two exposure variables (percent of overcrowdedness by ZCTA, and percent of multigenerational households by ZCTA); ZCTA-level prevalence (defined as a percentage) of coronary heart disease, obesity (defined as BMI ≥ 30kg/m^2^), and smoking; percent white; percent below the FPL; proportion of essential workers; and median income (defined in 2018 dollars). Coefficient results in the INLA model are presented with 95% Bayesian credible intervals. We assessed whether there was effect modification between the two exposures using an interaction term in the final fully adjusted model; the interaction term was not significant (β_interaction_ = 0.99, 95% credible interval: 0.99-1.00). Model outputs for all preceding models can be found in the Supplement.

The study protocol was approved by the [withheld for review] Institutional Review Board. All analyses were conducted in GeoDa version 1.16, QGIS version 3.16.2 and R version 4.0.3. Code to reproduce the analyses is available at: https://github.com/sara-venkatraman/NYC-Housing-COVID19.

## Results

### Descriptive Statistics

There were 39,923 suspected severe COVID-19 cases across 173 ZCTAs in NYC between March 1 and March 30, 2020. Figure 1 shows the proportion of overcrowded and multigenerational household in ZCTAs in NYC. The median proportion of overcrowded households by ZCTA was 5.06% (interquartile range [IQR] 4.68%), and the median proportion of multigenerational households by ZCTA was 6.88% (IQR 5.96%). Three of the top five ZCTAs in terms of cumulative suspected COVID-19 cases (11368, 11373, and 11208 – all found in the borough of Queens) were also in the top five ZCTAs in terms of percentage of overcrowdedness. The five ZCTAs with the highest proportion of multigenerational households were 11411, 11412, 11419, 11413, and 11429 – also found in Queens (see Figure 1).

**Figure 1:**
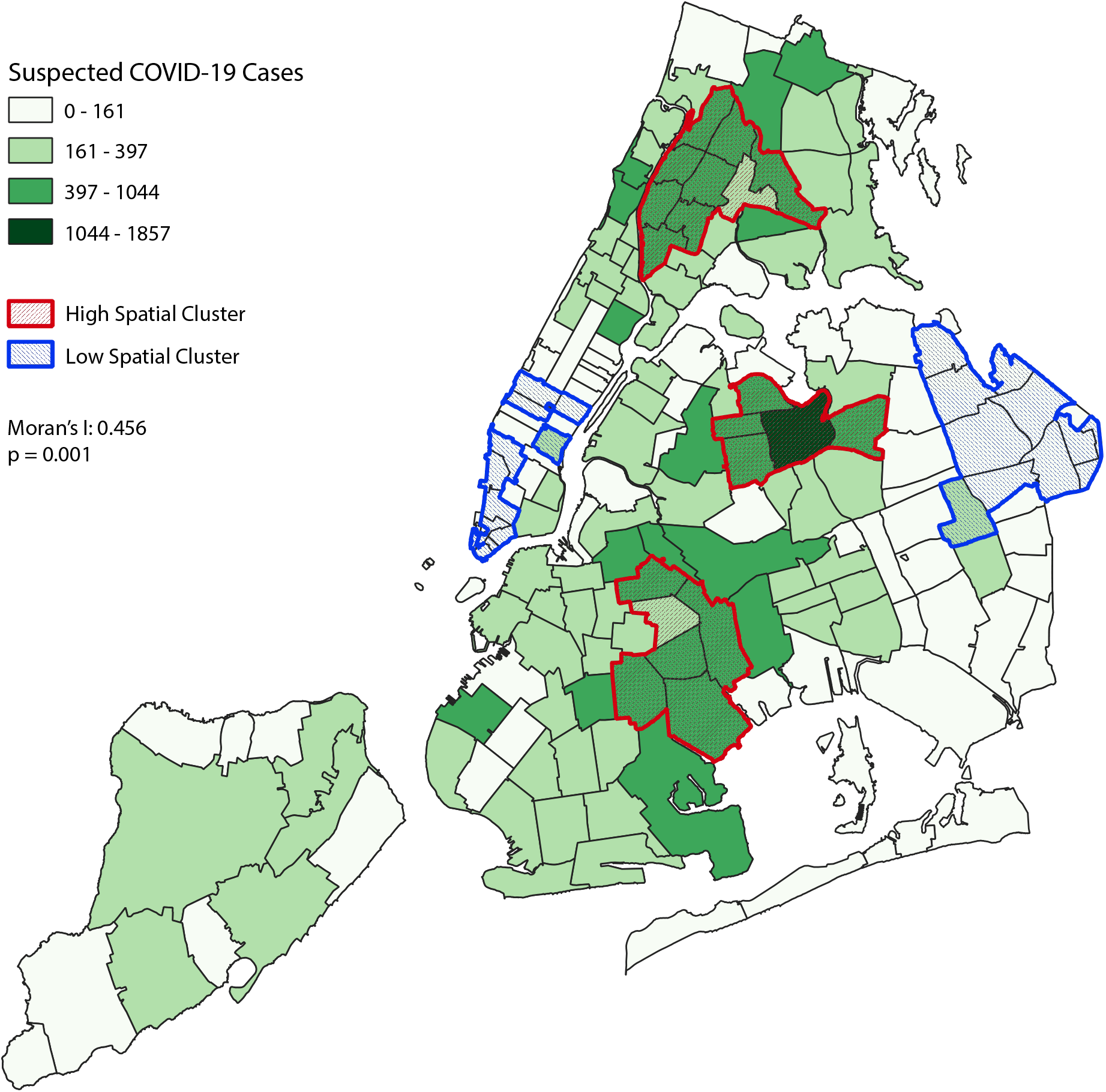
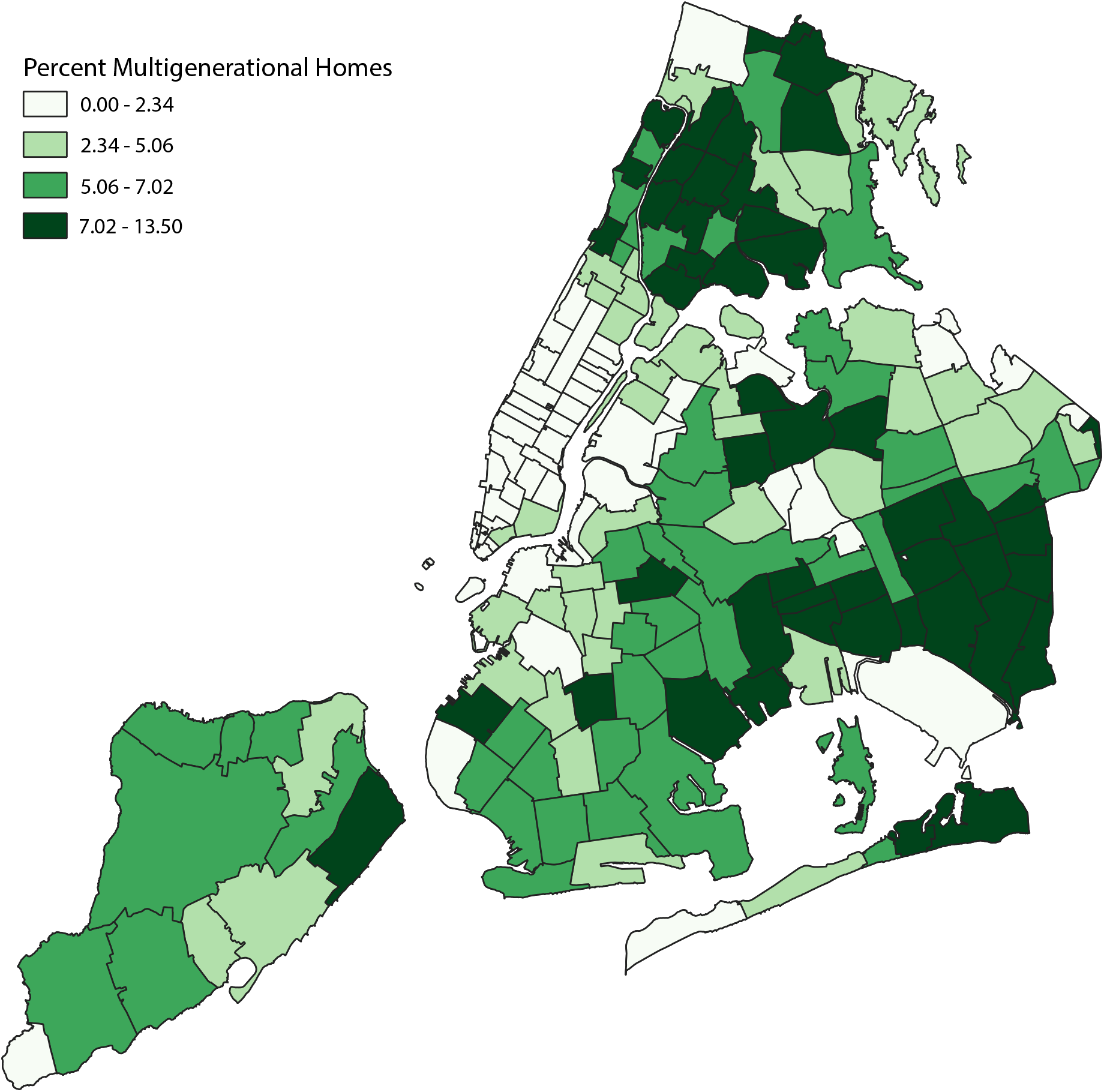
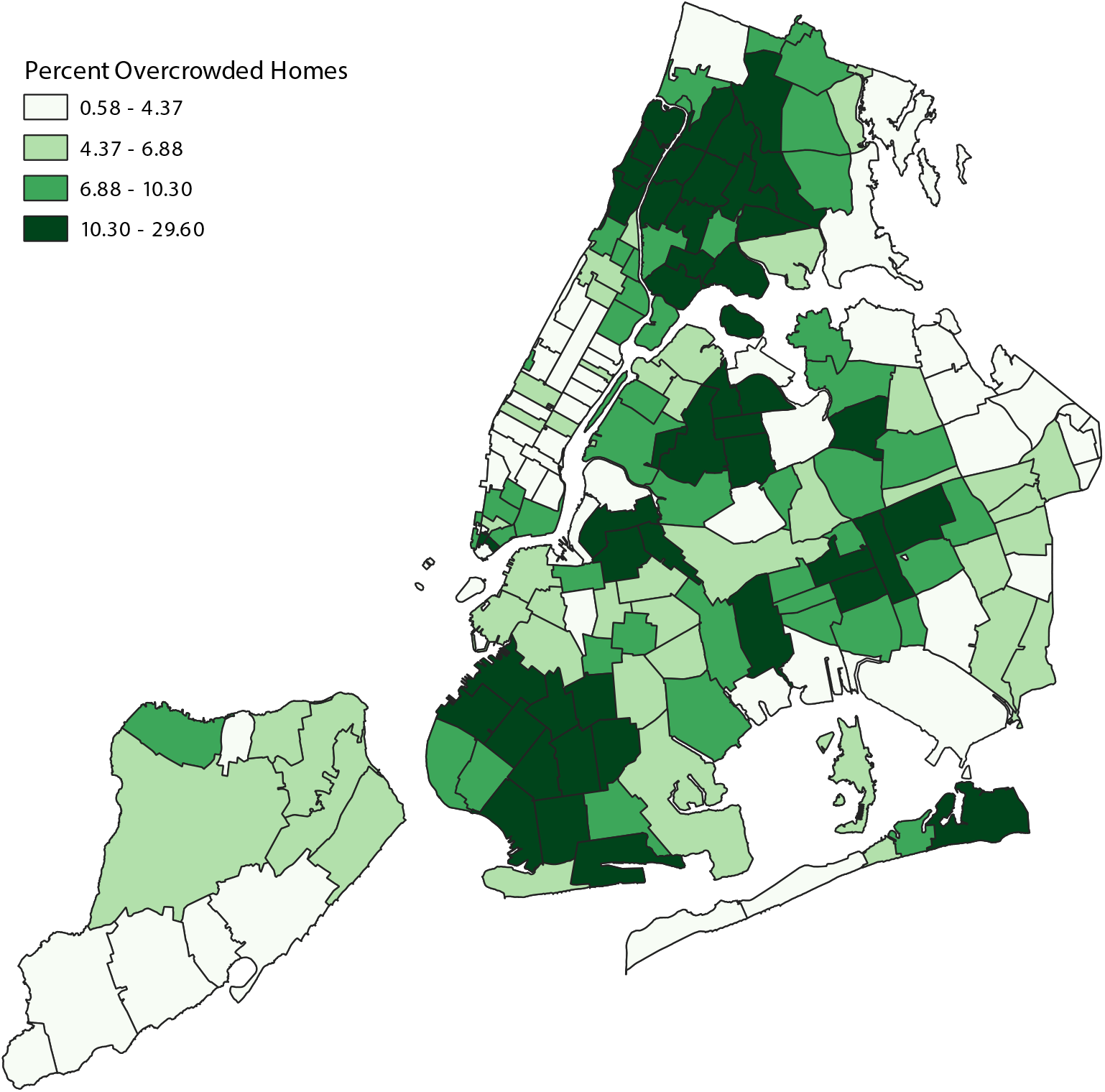
Distribution of New York City ZIP code tabulation area (ZCTA) by a) suspected COVID-19 cases in March 1-30 2020; and b) overcrowded housing^1^ and c) multigenerational housing^2^ in 2018. ^1^ Proportion of overcrowdedness defined as estimated number of housing units with more than one occupant per room, divided by the number of occupied housing units in each ZCTA, expressed as a percentage from 2018 American Community Survey 5-year estimates ^2^ Proportion of multigenerational housing defined as estimated percentage of residences occupied by grandparent and a grandchild less than 18 years of age in each ZCTA from 2018 American Community Survey 5-year estimates

Table 1 reports how, at a ZCTA-level, the prevalence of COVID-19 clinical risk factors and associated socioeconomic factors vary across quartiles of multigenerational households and overcrowded households. Compared to the first quartile, suspected severe COVID-19 cases rose almost four-fold in fourth quartile of ZCTAs with multigenerational households, and almost six-fold for overcrowded households. As the proportion of overcrowded households and multigenerational households rose by ZCTA in quartiles, there was a consistent increase in the prevalence of diabetes and smoking, as well as proportion of residents living below the FPL, while there was a consistent decline in the proportion of White residents and median income.

**Table 1:**
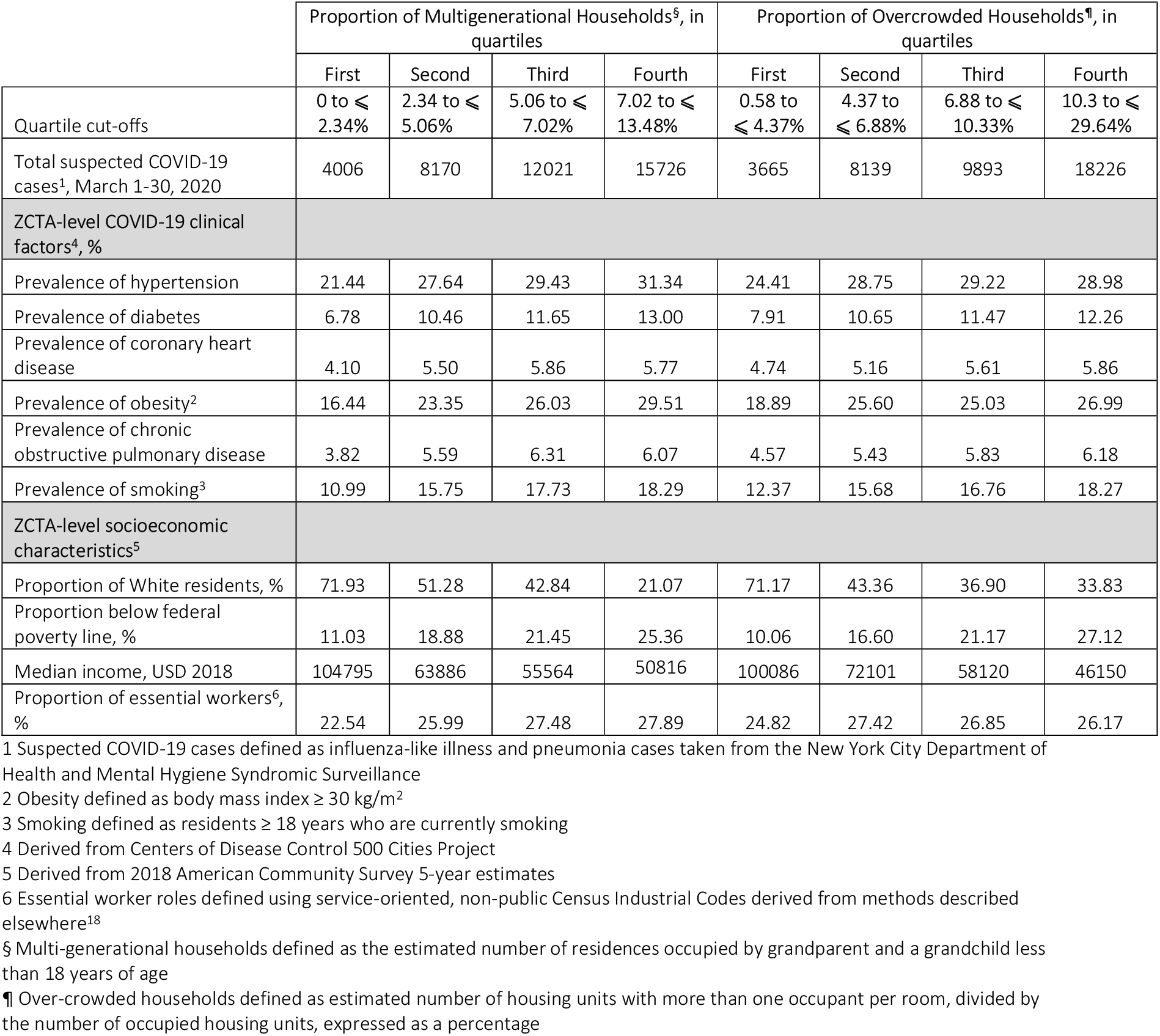
ZIP code tabulation area (ZCTA)-level clinical and socioeconomic characteristics by proportion of multigenerational households and proportion of overcrowded households, in quartiles

### Results from the Multivariable Models

Table 2 reports the results of unadjusted and fully adjusted analysis. Controlling for both COVID-19 clinical risk factors and socioeconomic characteristics which may increase risk of COVID-19 exposure, ZCTAs in the fourth quartile of percent overcrowdedness had a 67% increased risk of suspected severe COVID-19 cases compared to the first quartile (IRR 1.67, 95% CI = 1.12-2.52). Similarly, ZCTAs found in the fourth quartile of percent multigenerational households had a 77% increased risk of suspected severe COVID-19 cases compared to the first quartile (IRR 1.77, 95% CI = 1.11-2.79). Across both exposures, the case risk increased for each quartile, as noted in Figure 2A and 2B, which compare the observed suspected severe COVID-19 case rates with modelled rates.

**Table 2:** Adjusted incidence rate ratios (IRR) of suspected COVID cases per 10,000 for quartiles of ZIP code tabulation area-level (ZCTA)^1^.

|  | Unadjusted model |  | Fully adjusted model<sup>5</sup> |  |
| --- | --- | --- | --- | --- |
|  | IRR | 95% CI | IRR | 95% CI<sup>¶</sup> |
| **Percent of overcrowded households, in quartiles<sup>2</sup>**<br>**(ref: Quartile 1)** |  |  |  |  |
| Quartile 2 | 1.56 | 1.47 - 1.65 | 1.45 | 1.13 - 1.87 |
| Quartile 3 | 1.75 | 1.66 - 1.85 | 1.41 | 1.08 - 1.86 |
| Quartile 4 | 2.05 | 1.92 - 2.18 | 1.67 | 1.12 - 2.52 |
| **Percent of multigenerational households, in quartiles<sup>3</sup>**<br>**(ref: Quartile 1)** |  |  |  |  |
| Quartile 2 | 1.19 | 1.12 - 1.26 | 1.48 | 1.05 - 2.07 |
| Quartile 3 | 1.30 | 1.22 - 1.37 | 1.89 | 1.28 - 2.78 |
| Quartile 4 | 1.59 | 1.49 - 1.69 | 1.77 | 1.11 - 2.79 |
| **COVID-19 clinical risk factors, by ZCTA** |  |  |  |  |
| Prevalence of coronary heart disease |  |  | 0.92 | 0.82 - 1.03 |
| Prevalence of obesity (BMI ≥ 30kg/m<sup>2</sup>) | | | 0.97 | 0.94 - 0.99 |
| Prevalence of smoking |  |  | 1.08 | 0.99 - 1.17 |
| **Socioeconomic characteristics, by ZCTA** |  |  |  |  |
| Percent white |  |  | 0.99 | 0.99 - 1.00 |
| Percent below Federal Poverty Level |  |  | 0.97 | 0.95 - 0.99 |
| Median income, USD 2018 |  |  | 0.99 | 0.99 - 0.99 |
| Percent in defined essential worker role<sup>4</sup> |  |  | 1.01 | 0.98 - 1.04 |
| **Time, in days** | 1.04 | 1.04 - 1.05 | 1.04 | 1.04 - 1.04 |
| **Spatiotemporal structure accounted for?** | No |  | Yes |  |
| **Model family** | Generalized linear model |  | INLA<sup>6</sup> |  |

1 All covariates measured as percent at ZCTA-level; both models employ robust standard errors
2 Overcrowdedness defined as > 1 occupant per room in each household, derived from the 2018 American Community Survey 5-year estimates
3 Multi-generational households defined as the estimated number of residences occupied by a grandparent and a grandchild less than 18 years of age, derived from the 2018 American Community Survey 5-year estimates
4 Essential worker roles defined using service-oriented, non-public Census Industrial Codes derived from methods described elsewhere<sup>18</sup>
5 The fully adjusted model included percent of over-crowdedness and percent of multigenerational households simultaneously
6 INLA is integrated nested Laplace approximation method
¶ 95% Bayesian credible intervals

**Figure 2:**
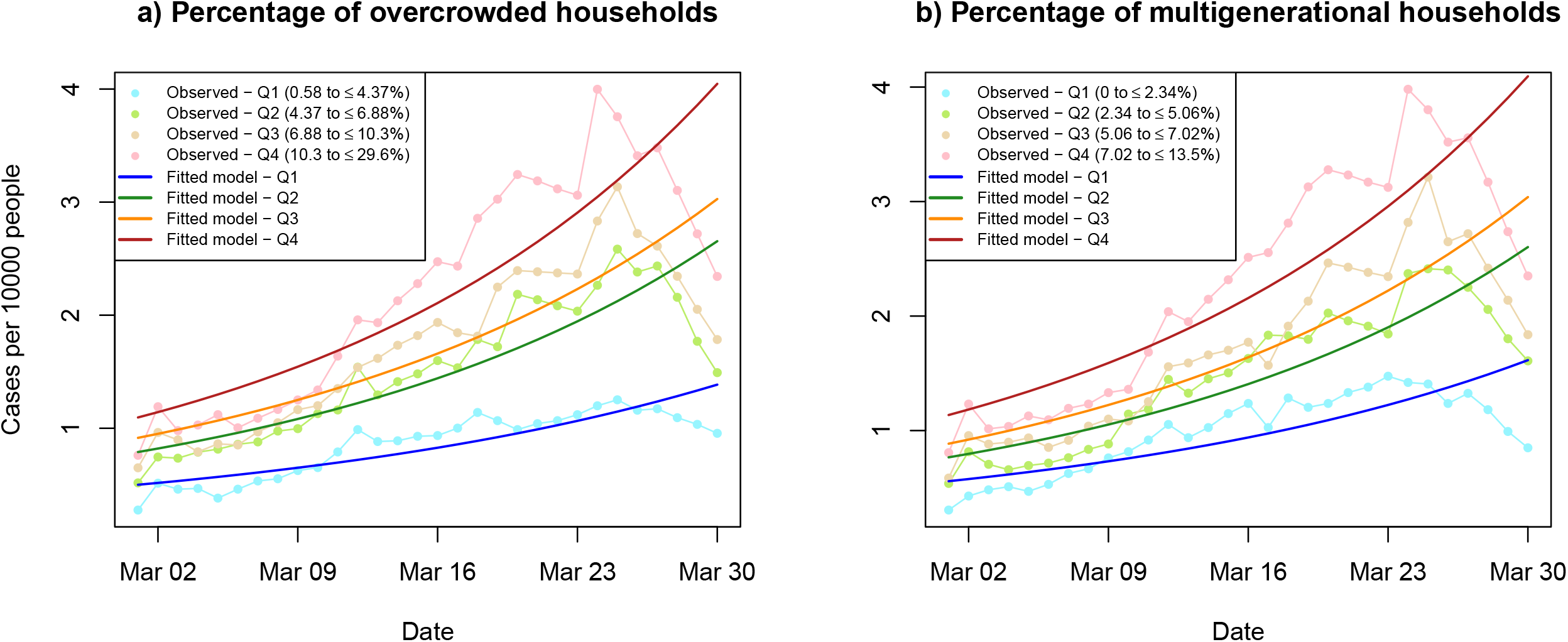
Unadjusted and predicted trends^1^ in suspected COVID-19 cases per 10,000 population in March 1-30 2020 by a) percentage of multigenerational households, and b) percentage of overcrowdedness, both in quartiles. ^1^ Predicted trends are taken from the fully adjusted model

## Discussion

In this ecological analysis of COVID-19 in NYC in March 2020, we found that ZCTAs with higher proportions of overcrowded and multigenerational households were associated with increased rates of suspected severe COVID-19 cases, after accounting for both socioeconomic factors which may increase the risk of infection with SARS-CoV-2, and clinical factors which may lead to more severe COVID-19 disease.

While several studies describe the role overcrowdedness may play as a risk factor for contracting COVID-19, this study is the first to establish the independent relationship between overcrowded households, multigenerational households, and suspected severe COVID-19 disease while adjusting for area-level socioeconomic and clinical characteristics. In a national dataset of county-level data, Ahmad et al. found that counties with higher proportions of poorer housing conditions (including over-crowding, high housing cost, or incomplete kitchen or plumbing facilities) had up to 50% higher risk of COVID-19 incidence in adjusted analyses.^12^ In NYC specifically, Emeruwa et al. found overcrowded housing to be an independent risk factor in a cohort of obstetric patients admitted for care during the COVID-19 surge.^12^ On the other hand, the role of multigenerational housing in the propagation of COVID-19 in the general population has been unclear. Anecdotal data exists,^19-21^ and the role of multigenerational housing has been suggested in the media as a potential accelerant in the spread of COVID-19, placing the elderly at risk.^22^ However, no study to date has quantitatively examined this potential risk factor. Since our analysis modeled both the proportions of overcrowdedness and multigenerational households simultaneously, our findings suggest that multigenerational households are independently associated with higher rates of suspected COVID-19 infection, and that this risk is independent of, and roughly equivalent to, that of overcrowding alone.

The results have notable scientific and clinical implications. First, by demonstrating a relationship between COVID-19 case rates and area-level measures of overcrowded and multigenerational housing, these findings may partly illustrate socioecological mechanisms that link observed inequities in COVID-19 related outcomes with the known biology of infectivity. Stark COVID-19-related racial/ethnic differences in mortality and morbidity exist, and data suggest that pre-hospitalization factors contribute to these differences.^1,23^ Although not addressed in our analysis, racial/ethnic differences in residential crowdedness and multigenerational households may play a role in COVID-19-related disparities in addition to that of essential workers. Since racial/ethnic and socioeconomic differences in multigenerational households exist, the association between multigenerational households and suspected COVID-19 case rates in our analysis may help to explain related disparities in COVID illness burden seen both in NYC and across the United States.^24^

Second, these results support recent developments in our understanding of the transmission dynamics of COVID-19. Our primary outcome, suspected severe COVID-19 cases defined as influenza-like illness or pneumonia, was captured at emergency departments throughout NYC. Although the health-seeking behavior of patients may have varied throughout the pandemic, patients presenting to the emergency department during the initial phases of the outbreak in NYC were likely to have had more severe symptomatic COVID-19 infections, which has been shown to be correlated to the concentration of inoculum received.^10,11^ These results add credence to the biologically plausible hypothesis of an association between transmission proximity, density of inoculum, and COVID-19 severity.

Third, our model demonstrates that certain commonly reported individual clinical and sociodemographic risk factors for COVID-19 hospitalization (e.g., presence of coronary heart disease,^25^ obesity,^16,26^ and minority status^1^) were not significantly associated with suspected COVID-19 cases at the area level. While the full meaning of this finding is unclear, it suggests that area-level risks that represent proximity in space, such as multigenerational housing and overcrowded households, may play a more powerful role in the spread of COVID-19 compared to individual clinical and sociodemographic COVID-19 risk factors. However, this is speculative and further investigation is warranted, ideally using hierarchical models of City-wide data containing both individual clinical and socioeconomic risk factors together with area-level measures which account for proximity in space.

Fourth, the location of the largest effects of overcrowdedness and multigenerational households on suspected COVID-19 cases reflects the on-the-ground reality of NYC’s initial COVID-19 wave.^27^ Although the study’s analysis was limited to the month of March, it became clearer as NYC’s cases rose through that month into early April that the boroughs of Queens and Brooklyn were inundated by COVID-19 cases.^28^ This reality corresponds to the spatial relationship between suspected severe COVID-19 cases and ZCTAs with higher proportions of multigenerational and overcrowded housing seen in our data.

## Limitations

Our study has several limitations. First, our analysis employed only area-level measures, which prevents making more generalizable statements at an individual level. However, our results concur with other studies that have employed area-level measures in their analysis, adding weight to our findings.^2,3^

Second, our analysis relied on a city-wide surveillance system tracking emergency room visits to determine suspected COVID-19 cases, which may not track with actual COVID-19 cases. Therefore, our dataset was likely to exclude both asymptomatic COVID-19 patients and those who chose not to present to emergency rooms (e.g., present to primary care physicians or urgent care facilities). Nevertheless, these data have been used in prior published reports to infer early COVID-19 case rates.^14^

Third, the surveillance data used may have included patients with non-COVID 19 respiratory illnesses such as influenza, which may have led to an overestimation the effect of overcrowdedness and multigenerational housing on COVID-19. However, analysis of emergency room utilization^29^ and anecdotal reports^30^ suggest that non-COVID-19 related patient volumes declined dramatically prior to the March 2020 NYC COVID-19 wave, due to both biological and social factors.

Finally, case rates may not accurately reflect the remaining population base of ZCTAs in NYC because of pandemic-related flight.^31^ Analysis of cell-phone data and mail forwarding requests comparing March 2 to May 1, 2020 has shown notable flight from particular neighborhoods throughout the borough of Manhattan, and those that abut the East River in the boroughs of Brooklyn and Queens. This may have reduced the total population at risk in our analysis, and led to underestimation of the neighborhood-level effects observed in our analysis. However, the neighborhoods that are the focus of this study were distinct from those reported to have the majority of pandemic flight, likely limiting the impact of this population shift on our results.

## Public Health Implications

COVID-19 lockdown policies globally have centered on some version of home quarantine, and/or school closure, leading to increased household “dwell time.” Though these policies have proved effective in the wider context, the implications of adherence to these measures for those living in close proximity to others, such as in overcrowded housing, have not been well explored.^32^ Our ecological analysis of the beginning of NYC’s spring COVID-19 wave suggests that lockdown-related school and non-essential business closures, which took place on March 13 and 23, 2020, were associated with adverse consequences for individuals living in areas characterized by high levels of household crowding and multigenerational households. Such public health measures may have amplified the household transmission of the SARS-CoV-2 virus and potentiated the severity of resulting COVID-19 cases. Since these housing characteristics follow racial/ethnic and socioeconomic divides, for example disproportionately affecting minority groups who serve as essential frontline workers, they may help explain resulting disparities in COVID-19 incidence. Future studies should seek to clarify this relationship to inform future public health interventions.

## Data Availability

Data is publicly available, and link to code provided in the manuscript

## Acknowledgements including declarations

This study received support from New York-Presbyterian Hospital (NYPH) and Weill Cornell Medical College (WCMC), including the Clinical and Translational Science Center (CTSC) (UL1 TR002384) and Joint Clinical Trials Office (JCTO)

This work was reviewed by the Weill Cornell Institutional Review Board

Dr. Ghosh is supported by National Center for Advancing Translational Sciences (NCATS) grant KL2-TR-002385 of the Clinical and Translational Science Center at Weill Cornell Medical College. Dr. Ibrahim is supported in part by a K24 Mid-Career Development Award from the National Institute of Arthritis and Musculoskeletal and Skin Diseases (K24AR055259).

Dr. Safford received salary support for investigator-initiated research from Amgen.

