## Supplement for "Association between overcrowded households, multigenerational households, and COVID-19: a cohort study"

**eTable 1: Adjusted incidence rate ratios (IRR) of suspected COVID cases per 10,000 for quartiles of ZIP code tabulation area-level (ZCTA)[1]**

| | Model 1 | | Model 2 | | Model 3 | | Model 4 | | Model 5 | |
|---|---|---|---|---|---|---|---|---|---|---|
| | IRR | 95% CI | IRR | 95% CI | IRR | 95% CI | IRR | 95% CI | IRR | 95% CI[¶] |
| **Percent of overcrowded households, in quartiles[2] (ref: Quartile 1)** | | | | | | | | | | |
| Quartile 2 | 1.56 | 1.47 - 1.65 | 1.22 | 1.15 - 1.30 | 1.36 | 1.28 - 1.45 | 1.22 | 1.14 - 1.29 | 1.45 | 1.13 - 1.87 |
| Quartile 3 | 1.75 | 1.66 - 1.85 | 1.47 | 1.39 - 1.56 | 1.43 | 1.34 - 1.52 | 1.44 | 1.35 - 1.54 | 1.41 | 1.08 -1.86 |
| Quartile 4 | 2.05 | 1.92 - 2.18 | 1.80 | 1.68 - 1.92 | 1.62 | 1.50 - 1.76 | 1.58 | 1.46 - 1.70 | 1.67 | 1.12 - 2.52 |
| **Percent of multigenerational households,[3] in quartiles (ref: Quartile 1)** | | | | | | | | | | |
| Quartile 2 | 1.19 | 1.12 - 1.26 | 1.08 | 1.00 - 1.15 | 1.01 | 0.94 - 1.08 | 1.17 | 1.09 - 1.26 | 1.48 | 1.05 - 2.07 |
| Quartile 3 | 1.30 | 1.22 - 1.37 | 1.09 | 1.01 - 1.18 | 1.08 | 1.00 - 1.16 | 1.38 | 1.27 - 1.50 | 1.89 | 1.28 - 2.78 |
| Quartile 4 | 1.59 | 1.49 - 1.69 | 1.03 | 0.93 - 1.12 | 1.27 | 1.15 - 1.39 | 1.54 | 1.40 - 1.70 | 1.77 | 1.11 - 2.79 |
| **COVID-19 clinical risk factors[4], by ZCTA (%)** | | | | | | | | | | |
| Prevalence of diabetes | | | 1.16 | 1.14 - 1.18 | | | | | | |
| Prevalence of coronary heart disease | | | 0.72 | 0.69 - 0.74 | | | 0.75 | 0.73 - 0.78 | 0.92 | 0.82 - 1.03 |
| Prevalence of obesity (BMI ≥ 30kg/m$^2$) | | | 1.01 | 1.01 - 1.02 | | | 1.03 | 1.03 - 1.04 | 0.97 | 0.94 - 0.99 |
| Prevalence of COPD[5] | | | | | | | | | | |
| Prevalence of smoking[6] | | | 0.99 | 0.98 - 1.01 | | | 0.98 | 0.96 - 0.99 | 1.08 | 0.99 - 1.17 |
| **Socioeconomic characteristics,[7] by ZCTA** | | | | | | | | | | |
| Percentage white | | | | | 0.99 | 0.99 - 1.00 | 0.99 | 0.99 - 1.00 | 0.99 | 0.99 - 1.00 |
| Percent below Federal Poverty Level | | | | | 0.99 | 0.98 - 0.99 | 0.97 | 0.97 - 0.98 | 0.97 | 0.95 - 0.99 |
| Median income, USD 2018 | | | | | 0.99 | 0.99 - 0.99 | 0.99 | 0.99 - 0.99 | 0.99 | 0.99 - 0.99 |
| Percentage in defined essential worker role[8] | | | | | 0.98 | 0.97 - 0.99 | 0.96 | 0.95 - 0.97 | 1.01 | 0.98 - 1.04 |
| **Time, days** | 1.04 | 1.04 -1.05 | 1.04 | 1.04 - 1.05 | 1.04 | 1.04 - 1.05 | 1.04 | 1.04 - 1.05 | 1.04 | 1.04 - 1.04 |
| **Space-time correlation matrix** | No | | No | | No | | No | | Yes | |
| **Model specification** | Generalized linear model | | Generalized linear model | | Generalized linear model | | Generalized linear model | | INLA[§] | |

1 All covariates measured as percent at ZCTA-level all models employ robust standard errors
2 Overcrowded households defined as > 1 occupant per room in each household, derived from 2018 American Community Survey 5-year estimates
3 Multigenerational households defined the estimated number of residences occupied by grandparent and a grandchild less than 18 years of age, derived from 2018 American Community Survey 5-year estimates
4 Derived from the Centers of Disease Control 500 Cities project
5 COPD is chronic obstructive pulmonary disease
6 Smoking defined as residents ≥ 18 years who are current smoking
7 Derived from 2018 American Community Survey 5-year estimates
8 Essential worker roles defined using service-oriented, non-public Census Industrial Codes derived from methods described elsewhere[18]
¶ 95% Bayesian credible intervals
§ INLA is integrated nested Laplace approximation
